# What Family Policies do Chinese Women Want? A Discrete Choice Experiment

**DOI:** 10.1101/2025.08.20.25333886

**Authors:** Yeqing Zhang, Liu Jue, Ana Correa, Jie Deng, Wenxin Yan, Stuart Gietel-Basten, Lu Gram

## Abstract

China’s fertility rate has been in continuous decline since 2013, despite policy shifts such as the relaxation of the one-child policy in 2015. The government is implementing a mix of family and work policies to support family formation, but little evidence exists on how Chinese women might respond. We conducted a single-profile discrete choice experiment (DCE) on a randomly selected sample of married women aged 21–49 from 31 provinces in mainland China. Participants were assigned hypothetical scenarios with varying levels of parental leave, childcare support, monetary incentives, and impacts on career progression. All policy attributes were significantly associated with intention to have a second child (p<0.001), with career progression having the largest effect. Compared to a two-year delay on promotions, leaving promotion prospects intact was associated with 5.0 percentage point increase in intention to have a second child. We estimated an implicit willingness-to-pay for career progression without penalties worth 743 CNY [104 USD] per month over three years, amounting to 26,763 CNY [3723 USD] in total. Our findings highlight the critical role of career progression in shaping decisions regarding family formation, pointing towards issues of gender equity at work as important barriers to reproductive aspirations.

## 1. INTRODUCTION

Projected demographic change is a major policy concern in China. The total population of China is forecast to decline from 1.42 billion in 2024 to 1.27 billion by 2050 and 638 billion by the end of the current century (UNDESA, 2024). China is also one of the most rapidly ageing countries on earth. In 2024, the population aged 20-59 was 817 million, while the population aged 60 and above was 292 million; a ratio of around 2.8:1. By 2050, these respective populations are forecast to be 589 million and 504 million; with the ratio forecast to be 1:1 by the end of the 2050s (UNDESA, 2024).

In common with many other countries around the world, these demographic changes have been accompanied with concerns among a wide range of policymakers and other actors (Gietel-Basten, 2023, Dorling and Gietel-Basten, 2017). In relation to social policy, concerns have been raised over the sustainability of existing health and social welfare systems, as well as the capacity to further enhance services such as long term care (Peng, 2021, Lopreite and Zhu, 2020, Peng and Yeandle, 2017, Bloom et al., 2015, Zhou et al., 2023). More broadly, these demographic concerns have been described as a threat to economic development, and ultimately, to geopolitical power (Wang et al., 2022, Coleman and Basten, 2015, Mitter, 2024, Stowe, 2005).

The government of China has, in response, prepared a wide array of policies to tackle both the causes and consequences of population ageing and decline. These policies are addressed in the 14th Five-Year Plan (2021–2025), a comprehensive framework the government routinely sets to guide national economic and social development. More specifically, Chapter 45 of the 14^th^ Five-Year Plan details ‘A National Strategy in Response to Population Aging’,^1^ outlining policies such as developing the silver economy, increasing the long-term care sectors, supporting at-home elderly care services and improving co-ordination of elderly care programs are proposed. In late 2024, the government also announced a long-awaited (if unpopular) reform in the retirement age (Zhang and Chand, 2024).^2^

A parallel set of policies, however, have been announced to directly target the declining fertility rate. China has a long history of direct population interventions, from Mao-era policies aiming at growing the population, to efforts to curb population growth from the late-1970s, i.e. the introduction of one-child policy in 1979. While there is much controversy over the direct impact of this policy on birth rates (Goodkind, 2017, Wang et al., 2018, Gietel-Basten et al., 2019), fertility decreased dramatically over the following decades.

In recent years, a marked shift from anti-natalism to pro-natalism in China has occurred (Wang et al., 2024). In 2013, the government lifted the one child restriction, allowing couples to have two children if one parent was an only child. This was expanded to a universal two-child policy in 2015 and further to a three-child policy in 2021 (Chen et al., 2023). To complement these changes, financial incentives were introduced in 2022, i.e. tax deductions of CNY 1,000 per month per child for children’s education or childcare under age three (Zhang et al., 2023). At the local/regional level, various context-specific incentive interventions have also been piloted in recent years, such as baby bonuses and tax breaks.^3^ Many such policies, however, focus only on the *direct* costs of childbearing, rather than the *indirect* or *opportunity* costs which are inevitably much higher for females (and their career progression) in the context of an ‘incomplete gender revolution’ of unequal sharing of caring and domestic responsibilities (Esping-Andersen, 2009, McDonald, 2000, McDonald, 2013).

The effectiveness and efficiency of such pronatalist policies in China remain uncertain. Despite the aforementioned policy interventions, fertility rates have continued to decline since 2013. Although the relaxation of the one-child policy in 2015 briefly boosted fertility rates, the effect was short-lived, with a sharp decline resuming from 2017 onward (Zhang et al., 2023). The Total Fertility Rate (TFR) fell from 1.88 in 2017 to 1.58 in 2018. By 2020, the TFR had dropped below the “lowest-low” fertility threshold of 1.3, and by 2022 it further declined to 1.08 (Zhai et al., 2023). National fertility surveys indicate a societal shift towards smaller family preferences, as most couples view having two children as the ideal and more than 20% prefer having only one child (Chen et al., 2023, Chen and Gietel-Basten, 2024).

Many scholars argue that a purely pronatalist response to population ageing (i.e. by increasing the fertility rate) is, in fact, inefficient and, in some cases, harmful (Gietel-Basten et al., 2022). For instance, newborns today will not enter the labour force until the 2040s, by which time the labour market demand will have changed and key challenges (e.g. the sustainability of the urban pension system) will have become almost unassailable. More broadly, evidence indicates that pronatalist policies based primarily on financial incentives are likely ineffective at significantly raising fertility rates (Gauthier and Gietel-Basten, 2024).

Nevertheless, recent policy pronouncements on family formation in China reveal a more nuanced picture, showing political will towards a more comprehensive and inclusive family policy regime. In the 14^th^ Five-Year Plan, ‘revised childbirth policies…to promote long-term and balanced population development’ were indeed included as part of the planned long-term population strategy. These more ‘inclusive’ policies are designed ‘in tandem with economic and social policies to reduce the burden of childbearing, upbringing, and education on families. Specific measures proposed include improving infant care, implementation of maternity leave policy, and exploring the possibility of mandating parental leave. In relation to maternal care, ‘a full range of services for maternal care, including preconception and postnatal care, and child development will be made available and easily accessible, to improve the health and wellbeing of the newborn population.’ More recently, State Council pronouncements have shifted its focus to ‘removing the barriers for individuals to meet their reproductive aspirations’ – something much more in line with international best practices as reflected in the Programme of Action of the International Conference on Population and Development – rather than top-down pronatalism.

China’s fertility support system, a network of policies and programs aimed at lowering barriers to childbearing and providing support for families, is still in its infancy. The initial attempts to address fertility barriers, such as the tax deduction introduced in 2022, have yet to see an effect on fertility rates (Dong et al., 2024, Zhang et al., 2023). More comprehensive and ambitious policy interventions are under-development and require further evidence-based justification. This study, therefore, aims to explore Chinese women’s preferences for family policies.

While family formation decisions are made at the household level, women’s preferences are crucial considering the currently unequal distribution of roles in childbearing and childcare (Doepke et al., 2023). Existing research identifies financial burdens, inadequate childcare support, and career disruption as primary concerns for women considering childbearing (Xiang et al., 2023, Yang et al., 2023, Liu et al., 2020). Historical progress in gender equity in education and employment has resulted in conflicts between women’s desires for self-actualisation and empowerment through work and traditional patriarchal and patrilineal family structures (Caldwell and Caldwell, 2005, Li and Jiang, 2019). Furthermore, rising childcare costs intersect with traditional Chinese expectations of intensive parental involvement in child-rearing, further complicating family formation decisions (Jones et al., 2009).

Various reviews have assessed the effectiveness of family policy interventions on outcomes such as maternal and child health and wellbeing, as well as on demographic measures such as the TFR (Gauthier and Gietel-Basten, 2024, Thévenon and Gauthier, 2011, Thévenon, 2011). These studies generally concluded that while such interventions (e.g. in childcare, parental care, and financial support to families) can improve outcomes for parents and children, they have limited impact on the overall TFR without either substantial financial investment and/or cultural change.

The majority of these reviews, however, have focussed on OECD and/or European settings. Cultural differences and national contexts are critical to understanding the potential effects of any family policy intervention. Given the short history of family policy interventions in China, it is not yet feasible to perform a systematic evaluation of such policies. It is, however, possible to explore *prospectively* which policies might have the greatest impact on reducing barriers to childbearing in China.

In this study, we conducted a Discrete Choice Experiment (DCE) using a nationally representative online survey. Our analysis focused on a subsample of married women with one child, examining their intentions to have a second child - a choice that remains infrequent despite the relaxation of China’s birth control policies since 2013 but crucial for maintaining the population replacement rate (Chen et al., 2023). Moreover, women with firsthand experience of childbearing may be better positioned to provide realistic evaluations of support needed for motherhood, compared to women who have never had children before. Our study contributes to existing literature by exploring prospectively how various family support policies, targeting both direct and indirect cost of child bearing, would affect women’s fertility intention in China.

## 2. METHODS

### 2.1 Design Overview

We conducted a DCE, a commonly used stated-preference method in healthcare research (Clark et al., 2014), to understand how hypothetical family policies might influence women’s intentions to have a second child. This method involves presenting survey respondents with a series of hypothetical scenarios, each representing a set of policies with varying levels of support, and asking them to indicate their fertility intentions for each scenario. The variation in respondents’ choices enables us to estimate how they value different polices.

### 2.2 Design of the Single Profile DCE

#### 2.2.1 Attributes

The DCE tool was designed to have five attributes: maternity leave, paternity leave, childcare support, monetary incentives, and career impact. Attributes were selected based on a rapid review of fertility barriers and family policies worldwide, followed by a consultation with a government fertility policy expert (author JL). Policies targeting financial burdens, childcare support, and work-family conflict emerged as the most relevant. Specific levels of each policy attribute were informed by existing policies in China and from other low-fertility contexts, including OECD and East Asian countries (Choi et al., 2018, Shen et al., 2020, Jones, 2019). The attribute levels were further refined after conducting a pilot study with 20 respondents. The selected attributes, along with their levels are summarised in Table 1. See Appendix A for a detailed justification of each attribute and level.

**Table 1:** Attribute and levels for the Discrete Choice Experiment.

| Attributes | Levels |
| --- | --- |
| Maternity Leave | 1 - 98 days' fully paid maternity leave<br>2 - 158 days' fully paid maternity leave<br>3 - 270 days' fully paid maternity leave |
| Paternity Leave | 1 - No fully paid paternity leave<br>2 - 20 days' fully paid paternity leave<br>3 - 98 days' fully paid paternity leave |
| Childcare Support | 1 - No free access to crèche<br>2 - Free access to crèche 2 days a week for the first three years<br>3 - Free access to crèche 5 days a week for the first three years |
| Incentives | 1 - No<br>2 - 500 CNY childcare subsidy per month the first three years<br>3 - 1000 CNY childcare subsidy per month the first three years<br>4 - 1500 CNY childcare subsidy per month the first three years |
| Career Impact | 1 - Promotion delayed by two years due to having a baby<br>2 - Promotion delayed by one year due to having a baby<br>3 - Promotion not affected |

#### 2.2.2 Tasks

We adopted a single-profile DCE approach, where respondents evaluate one option at a time rather than choosing between two. We chose this design, because we intended to evaluate the impact of policy packages on fertility intentions, which necessarily requires comparison of (1) a package of fertility policies plus another child with (2) the option of not having another child (in which case none of the family policies apply). Comparing multiple packages of family policies would only allow us to elicit participants’ policy preferences *conditional* on having already decided to have another child, which was not the aim of this study. Furthermore, single-profile DCEs have in fact been shown to better simulate real-world decision-making, reduce cognitive load for participants, and yield informed and reliable answers compared to traditional two-profile DCE designs (Bridges et al., 2011, Díaz Luévano et al., 2021).

The experiment consisted of a series of hypothetical scenarios, each presenting participants with a unique combination of five policy attributes at different levels. Each scenario required a forced-choice response, which maximises information gained and minimises biases that could be introduced by confounders associated with both fertility intentions and opt-out behaviours (Janssen et al., 2018). Participants were asked how strongly they would prefer having a second child under the given hypothetical policy package. A 6-point Likert scale was used for responses ranging from “absolutely does not want a second child” to “absolutely want a second child.” An example of the scenario format is provided in Table 2.

**Table 2:** Example DCE question.

|  |  |
| --- | --- |
| Maternity Leave<br>(fully paid) | 158 days |
| Paternity Leave<br>(fully paid) | 20 days |
| Free creche services<br>(for child under 3) | 2 days per week |
| Childcare subsidies<br>(for child under 3) | 500 CNY per month |
| Promotion | Delayed by 1 year |

Given the above policies, to what extent would you like to have another child **in the next 3 years**?
Absolutely don't want
Absolutely want

A full-factorial design that includes all possible combinations of the five attributes across their different levels yields 3^4 × 4 = 324 unique profiles (West and Ramcharan, 2019). However, some profiles are dominated by others, and similar profiles may add limited value. To enhance efficiency, we applied a D-efficiency criterion and selected 48 orthogonal and balanced profiles, which ensures minimal loss of information while accommodates respondent heterogeneity (Bridges et al., 2011). To further reduce cognitive overload, the 48 profiles were randomly blocked into four survey versions-each containing 12 scenarios. Respondents were randomly assigned to one of the four.

The DCE tool was pre-tested with a sample of 20 women varying in age, educational attainment, and occupation. During the pre-testing phase, each respondent was interviewed to assess their understanding of the questions and the relevance of the attributes to their fertility decision-making. Designs were refined accordingly. We estimated that a minimum sample size of 665 participants was required to achieve the desired statistical power (1 - *β* = 0.8) at a 5% significance level for each policy attribute, using methods proposed by De Bekker-Grob et al. (2015). Additionally, the ISPOR guidelines show a sample size of 500 as a practical balance between precision gains and cost constraints (Bridges et al., 2011).

### 2.3 Data Collection

#### 2.3.1 Setting and Respondents

A cross-sectional survey was conducted across 31 provinces in China from 8 July 2024 to 22 August 2024. An online questionnaire was distributed via Wenjuanxing (https://www.wjx.cn/), one of the largest online survey platforms in the country, which maintains a nationally representative sample database comprising more than 6.2 million registered users and 10 million daily active users. Participants were recruited via web, email, or text messages from the sample pool of Wenjuanxing. Participants who were female, married, and of reproductive age (21-49 years) were eligible for inclusion in the study. Sampling was conducted proportional to the total number of married women in each province in China and to the population size of the married woman within each age group: 21–30, 31– 40, and 41–50.

A total of 2,559 married women of fertile age were contacted, and 2,146 (83.9%) consented to participate and completed the questionnaire. During data validation, respondents who provided inconsistent answers to duplicated test questions and logically related questions, were excluded. After validation, we were left with a complete sample of 2,032 respondents. Their geographic distribution is shown in Figure 1, where darker shades indicate higher proportions of the total sample.

**Figure 1:**
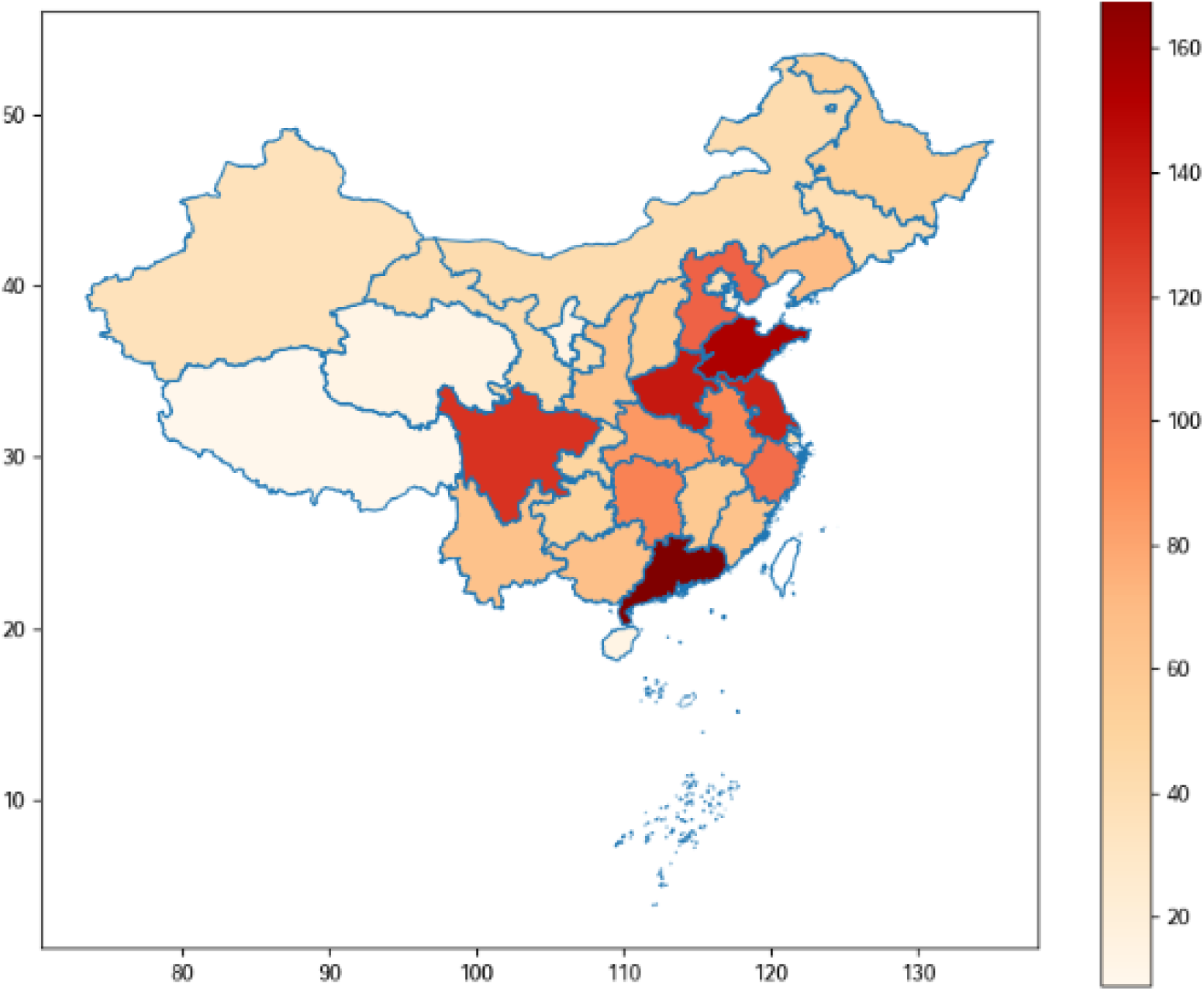
Geographic distribution of the study sample

As the study focused on women’s intentions for a second child, respondents with no children or more than one child were excluded, along with other groups, such as LGBTQ+ couples and individuals unable to conceive. The final sample for the main analysis consisted of 1,005 participants. For sensitivity analysis, similar models were tested on different samples: women considering whether to have their first child (n=230) and those considering a third child (n=432).

#### 2.3.2 Measures

In addition to the DCE tasks, the survey collected data on respondents’ sociodemographic characteristics and the transgenerational childcare support they have. We also assessed respondents’ fertility intentions in their actual life, before presenting the DCE section, with the question “Do you plan on having another child within the next three years?”

### 2.4 Data Analysis

Our primary analysis estimated married women’s preferences for family policies using a mixed ordinal logistic regression. First, we modelled respondent’s utility of having another child in any DCE scenarios as a continuous variable, varying linearly with policy attributes. It follows the following specification:

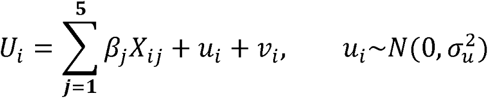

Here, *x_ij_* represents policy attributes and *β_j_* are the corresponding preference weights, assumed to be constant across individuals. Monetary incentives were represented as continuous variables, while all other attributes were modelled as categorical variables. *u_i_* is an individual-level random effect introduced to capture variation in individual preferences. *υ_i_* is an error term assumed to follow a type 1 extreme value distribution.

Following the principles of random utility maximisation theory (Dell’Olio et al., 2018), we assumed that respondents’ choices of the strength of their fertility intension in the DEC scenario, in discreet levels from 0 to 5, were driven by the utility they gained. A mixed ordinal logistic regression was utilised to map higher utility values to higher categories of fertility intention. The method accounts for the ordered nature of the response variable without imposing the restrictive assumption of equal utility gains across intention categories by use of ordinary linear regression model. We further accounted for individual’s preference heterogeneity by including a random intercept *u_i_*. For all attribute levels, odds ratios (ORs) with 95% confidence intervals were reported as preference weight measures. Marginal effects of policy change on fertility intentions were also calculated to show the additional probability gains for leaning towards higher intentions using the delta-method.

We calculated willingness to pay (WTP) for differing policy levels. To improve precision and incorporate uncertainty, we used simulations to derive WTP distributions by dividing draws from the distributions of non-monetary coefficients by draws from the distribution of monetary incentive coefficient using the bootstrapping method.

In our heterogeneity analysis, we explored whether women’s policy preferences differed across intention levels, using a partial proportional odds (PPO) model, following Williams (2016). We evaluated the interaction effects between policy attributes, as well. as between policy attributes and individual sociodemographic characteristics, such as annual household income. We explored whether women’s preferences for family formation support policies differed when considering having a first, second, or third child. Finally, we conducted sensitivity analysis to accommodate decision inertia, a phenomenon in which choices made in previous DCE scenarios influence choices in subsequent scenarios.

For further methodological details of the above analyses, including equations providing full model specifications, please see Supplementary Appendices A-D. Weighting was used in all the analyses to accommodate the attrition during data cleaning and improve the representativeness of married women in each province of China. Data analysis was performed in Stata, version 18.5.

### 2.5 Ethics

This study received approval from the UCL Research Ethics Committee (ANONYMISED FOR REVIEW) and Peking University Third Hospital Medical Science Research Ethics Committee (ANONYMISED FOR REVIEW).

## 3. RESULTS

### 3.1 Descriptive Statistics

A sample of 1,005 married women with one child in China was used for analysis. Table 3 summarises the descriptive statistics and presents demographic and socioeconomic differences between those planning to have a second child within the next three years and those not planning. Demographic profiles between the two groups were largely similar, except that respondents in the group planning to have a second child were younger (32% aged 21–30; 14% aged 41-50) than the group not planning to do so (19% aged 21–30; 36% aged 41-50). Annual household income was also significantly higher (40% reporting incomes of 400,000 CNY or more when planning to have a second child compared to only 24% when not planning to do so). Support from grandparents was common across the sample, with 59% receiving childcare support at least five days a week. Such support was more frequent in the group planning to have a second child, with 40% receiving daily assistance compared to 27% in the other group.

**Table 3:**
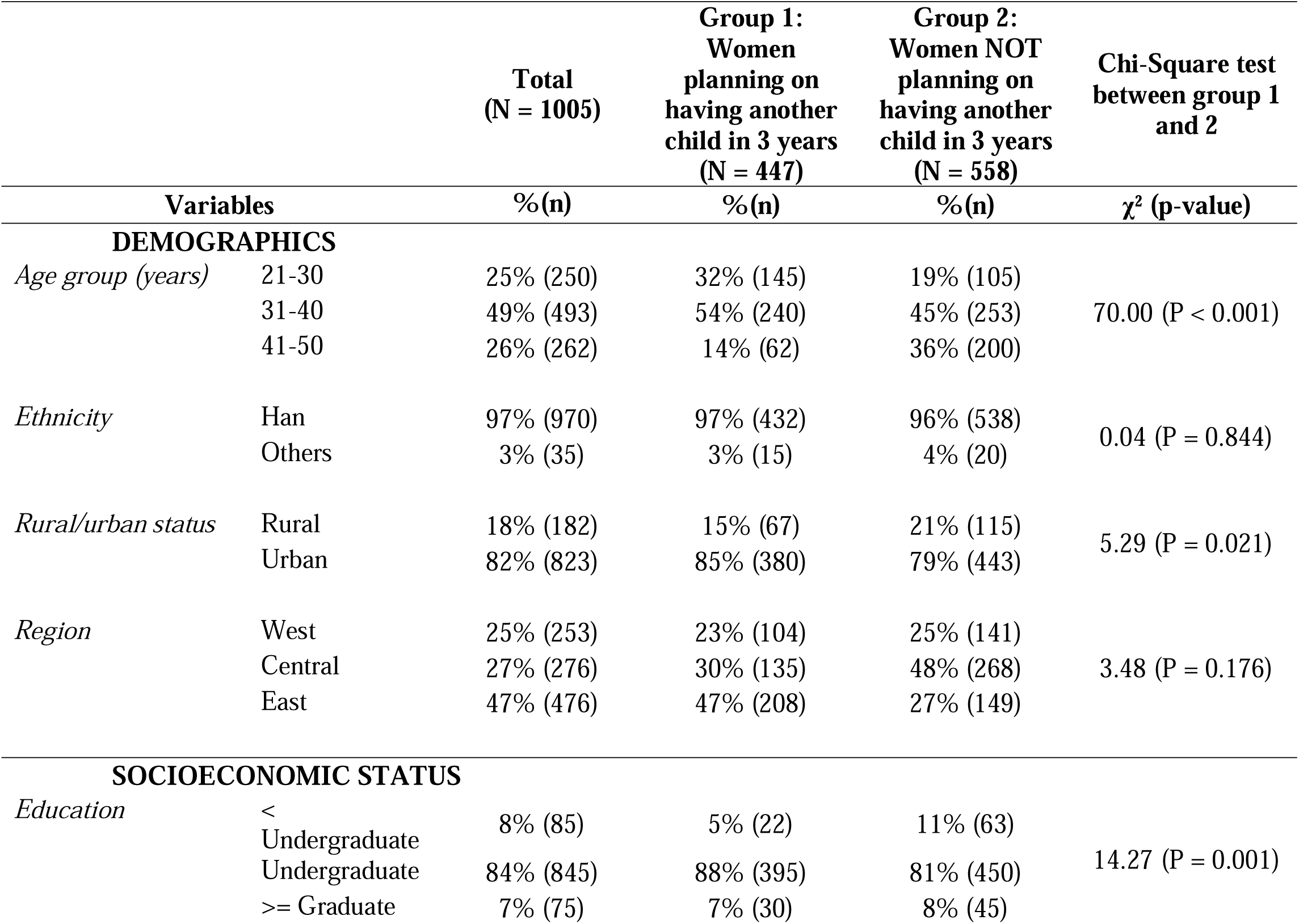
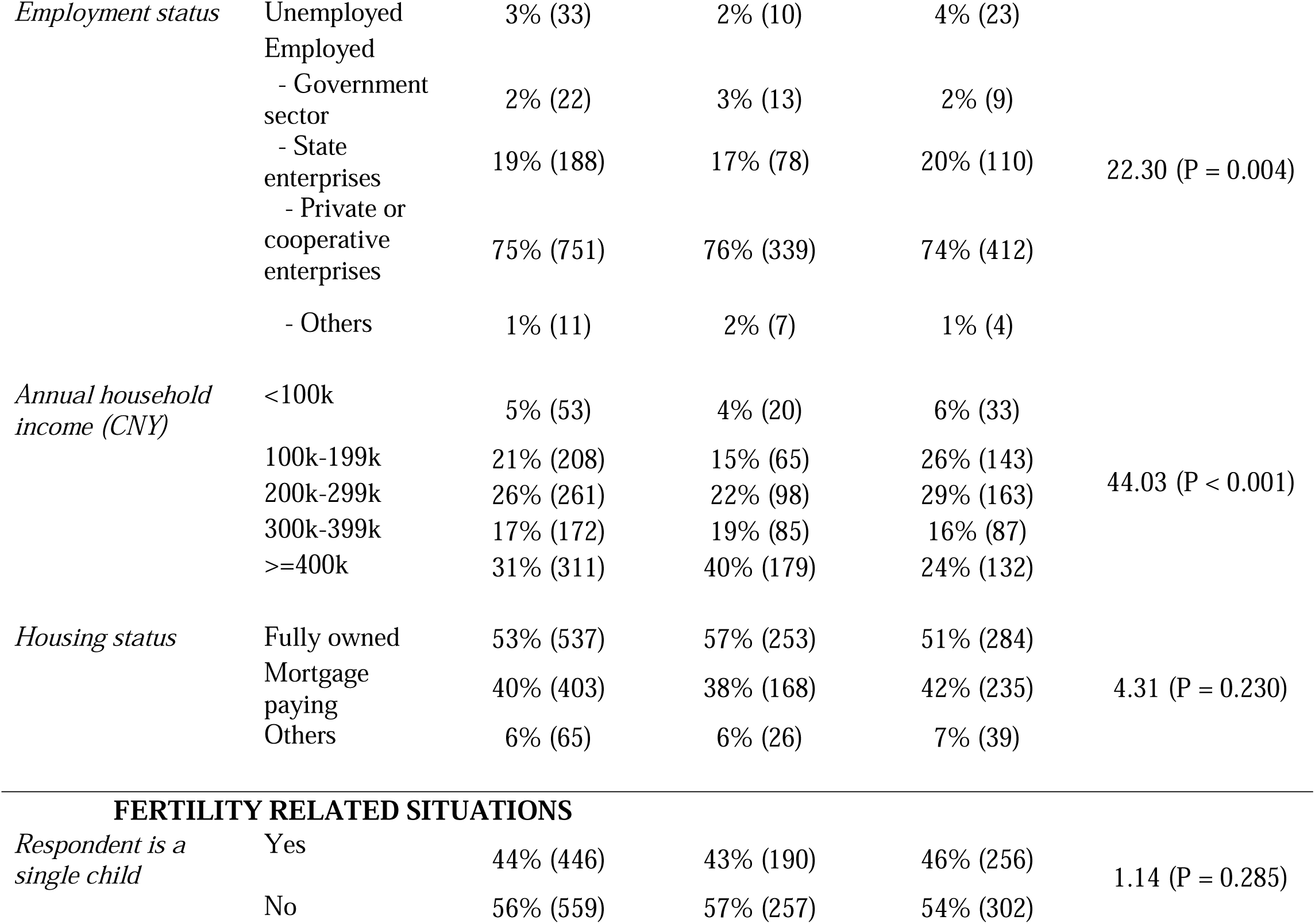
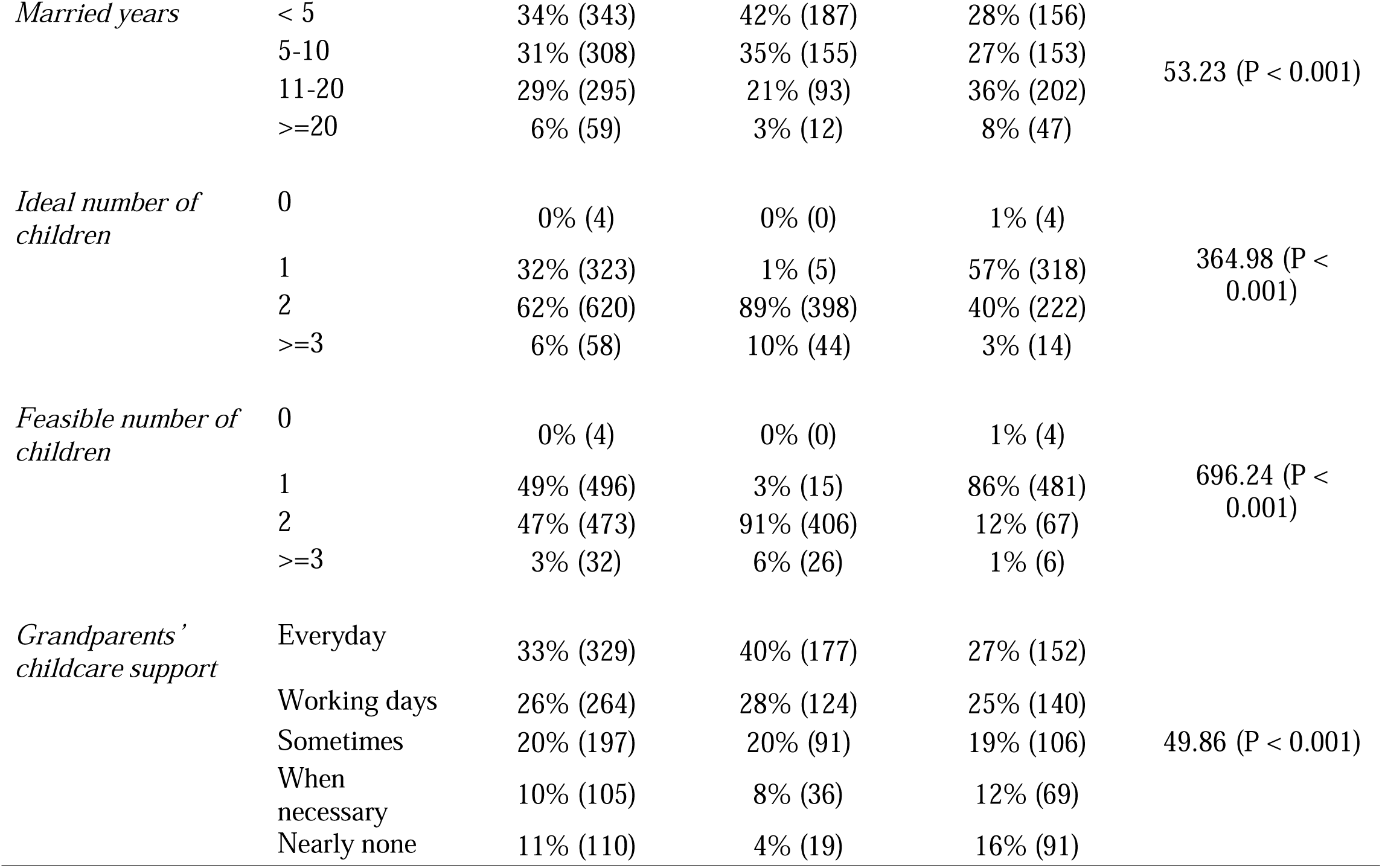
Descriptive statistics of respondents.

|  |  | Total<br>(N = 1005) | Group 1:<br>Women<br>planning on<br>having another<br>child in 3 years<br>(N = 447) | Group 2:<br>Women NOT<br>planning on<br>having another<br>child in 3 years<br>(N = 558) | Chi-Square test<br>between group 1<br>and 2 |
| --- | --- | --- | --- | --- | --- |
| Variables | | %(n) | %(n) | %(n) | $\chi^2$ (p-value) |
| DEMOGRAPHICS |  |  |  |  |  |
| Age group (years) | 21-30 | 25% (250) | 32% (145) | 19% (105) | 70.00 (P < 0.001) |
|  | 31-40 | 49% (493) | 54% (240) | 45% (253) |  |
|  | 41-50 | 26% (262) | 14% (62) | 36% (200) |  |
| Ethnicity | Han | 97% (970) | 97% (432) | 96% (538) | 0.04 (P = 0.844) |
|  | Others | 3% (35) | 3% (15) | 4% (20) |  |
| Rural/urban status | Rural | 18% (182) | 15% (67) | 21% (115) | 5.29 (P = 0.021) |
|  | Urban | 82% (823) | 85% (380) | 79% (443) |  |
| Region | West | 25% (253) | 23% (104) | 25% (141) | 3.48 (P = 0.176) |
|  | Central | 27% (276) | 30% (135) | 48% (268) |  |
|  | East | 47% (476) | 47% (208) | 27% (149) |  |
| SOCIOECONOMIC STATUS |  |  |  |  |  |
| Education | < Undergraduate | 8% (85) | 5% (22) | 11% (63) | 14.27 (P = 0.001) |
|  | Undergraduate | 84% (845) | 88% (395) | 81% (450) |  |
|  | >= Graduate | 7% (75) | 7% (30) | 8% (45) |  |
| *Employment status* | Unemployed | 3% (33) | 2% (10) | 4% (23) | 22.30 (P = 0.004) |
|  | Employed |  |  |  |  |
|  | - Government sector | 2% (22) | 3% (13) | 2% (9) |  |
|  | - State enterprises | 19% (188) | 17% (78) | 20% (110) |  |
|  | - Private or cooperative enterprises | 75% (751) | 76% (339) | 74% (412) |  |
|  | - Others | 1% (11) | 2% (7) | 1% (4) |  |
| *Annual household income (CNY)* | <100k | 5% (53) | 4% (20) | 6% (33) | 44.03 (P < 0.001) |
|  | 100k-199k | 21% (208) | 15% (65) | 26% (143) |  |
|  | 200k-299k | 26% (261) | 22% (98) | 29% (163) |  |
|  | 300k-399k | 17% (172) | 19% (85) | 16% (87) |  |
|  | >=400k | 31% (311) | 40% (179) | 24% (132) |  |
| *Housing status* | Fully owned | 53% (537) | 57% (253) | 51% (284) | 4.31 (P = 0.230) |
|  | Mortgage paying | 40% (403) | 38% (168) | 42% (235) |  |
|  | Others | 6% (65) | 6% (26) | 7% (39) |  |
| FERTILITY RELATED SITUATIONS |  |  |  |  |  |
| *Respondent is a single child* | Yes | 44% (446) | 43% (190) | 46% (256) | 1.14 (P = 0.285) |
|  | No | 56% (559) | 57% (257) | 54% (302) |  |
| *Married years* | < 5 | 34% (343) | 42% (187) | 28% (156) | 53.23 (P < 0.001) |
|  | 5-10 | 31% (308) | 35% (155) | 27% (153) |  |
|  | 11-20 | 29% (295) | 21% (93) | 36% (202) |  |
|  | >=20 | 6% (59) | 3% (12) | 8% (47) |  |
| *Ideal number of children* | 0 | 0% (4) | 0% (0) | 1% (4) | 364.98 (P < 0.001) |
|  | 1 | 32% (323) | 1% (5) | 57% (318) |  |
|  | 2 | 62% (620) | 89% (398) | 40% (222) |  |
|  | >=3 | 6% (58) | 10% (44) | 3% (14) |  |
| *Feasible number of children* | 0 | 0% (4) | 0% (0) | 1% (4) | 696.24 (P < 0.001) |
|  | 1 | 49% (496) | 3% (15) | 86% (481) |  |
|  | 2 | 47% (473) | 91% (406) | 12% (67) |  |
|  | >=3 | 3% (32) | 6% (26) | 1% (6) |  |
| *Grandparents' childcare support* | Everyday | 33% (329) | 40% (177) | 27% (152) | 49.86 (P < 0.001) |
|  | Working days | 26% (264) | 28% (124) | 25% (140) |  |
|  | Sometimes | 20% (197) | 20% (91) | 19% (106) |  |
|  | When necessary | 10% (105) | 8% (36) | 12% (69) |  |
|  | Nearly none | 11% (110) | 4% (19) | 16% (91) |  |

### 3.2 Preference Weights

Table 4 presents estimated preference weights for the five family formation support policies. All attributes were significantly associated with intention to have a second child (p<0.001). Career progression showed the largest effect, followed by paid maternity and paternity leave, childcare support, and monetary incentives. Compared to a two-year delay in career progression due to childbearing, having no delay was associated with 4.75 times higher odds of choosing categories representing stronger second-child intentions (p<0.001, 95% CI 4.27 to 5.28), whereas a one-year delay was associated with a 1.54 times increase in odds (p<0.001, 95% CI 1.40 to 1.70).

**Table 4:** Preference weights and willingness to pay (WTP)

|  | Preference Weights |  | WTP |  |
| --- | --- | --- | --- | --- |
|  | OR | 95% CI | Unit: 100s<br>of CNY | 95% CI |
| ***Baby stipend (effect in 100s of CNY)*** | 1.23*** | [1.22,1.25] | - | - |
| ***Paid maternity leave*** |  |  |  |  |
| 98 days |  | *Reference Level* |  |  |
| 158 days | 1.92*** | [1.75,2.10] | 3.10 | [2.58, 3.63] |
| 270 days | 3.93*** | [3.49,4.43] | 6.53 | [5.75, 7.32] |
| ***Paid paternity leave*** |  |  |  |  |
| None |  | *Reference Level* |  |  |
| 20 days | 2.59*** | [2.36,2.83] | 4.53 | [3.99, 5.07] |
| 98 days | 3.47*** | [3.14,3.83] | 5.93 | [5.21, 6.66] |
| ***Childcare support*** |  |  |  |  |
| None |  | *Reference Level* |  |  |
| Free access to crèche 2 days a week for 3 years | 1.62*** | [1.48,1.77] | 2.29 | [1.83, 2.74] |
| Free access to crèche 5 days a week for 3 years | 2.54*** | [2.25,2.86] | 4.44 | [3.86, 5.02] |
| ***Career progression*** |  |  |  |  |
| Promotion delayed by 2 years |  | *Reference Level* |  |  |
| Promotion delayed by 1 year | 1.54*** | [1.40,1.70] | 2.07 | [1.63, 2.50] |
| Promotion not affected | 4.75*** | [4.27,5.28] | 7.43 | [6.73, 8.14] |

OR = Odds Ratio, CI = Confidence Interval
\* $p < 0.05$ , \*\* $p < 0.01$ , \*\*\* $p < 0.001$ **Table 5:** Marginal effects of choosing categories representing lowest and highest

Extending maternity leave from 98 to 158 days increased the odds of having higher intention for a second child by 1.92 times (p<0.001, 95% CI 1.75–2.10), while extending it to 270 days further raised the factor to 3.93 times (p<0.001, 95% CI 3.49–4.43). Paternity leave exhibited a less pronounced effect: an extension from 0 to 20 days increased the odds by 2.59 times (95% CI 2.36 to 2.83), with a smaller gain to 3.47 times if extending to 98 days (p<0.001, 95% CI 3.14 to 3.83).

Providing free crèche access two days a week was associated with 1.62 times higher odds of favouring a second child (p<0.001, 95% CI 1.48 to 1.77), increasing to 2.54 times with full-time access (p<0.001, 95% CI 2.25 to 2.86) as compared to no free access.

For monetary incentives, every additionally 100 CNY provided in monthly baby stipends over three years was associated with 1.23 times higher odds of favouring a second child (p<0.001, 95% CI 1.22 to 1.25).

### 3.3 Willingness-to-pay

The WTP estimates in the right-hand panel of Table 4 illustrate the value respondents placed on each family policy. In general, respondent WTP aligned with preference weights. Respondents demonstrated the highest WTP for career progression without penalties, which constituted 743 CNY [*103 USD* ^4^] per month (95% CI 673 to 814) over three years, amounting to 26,763 CNY [*3,723 USD*] in total. WTP for reducing career delay from two years to one was 207 CNY [*29 USD*] per month (95% CI 163 to 250), totalling 7,437 CNY [*518 USD*] over three years.

For parental leave durations, respondents showed a monthly WTP of 453 CNY [*49 USD*] (95% CI 399 to 507) for a 20-day paid paternity leave and 593 CNY [*65 USD*] (95% CI 521 to 666) for a 98-day paid paternity; a monthly WTP of 310 CNY [*34 USD*] (95% CI 258 to 363) for a 60-day extension of maternity leave and 653 CNY [*71 USD*] (95% CI 575 to 732) for a 172-day extension of maternity leave.

Regarding childcare services, respondents’ monthly WTP was 229 CNY [*32 USD*] (95% CI 183 to 274) for part-time access and 444 CNY [*48 USD*] (95% CI 386 to 502) for full-time access.

### 3.4 Predicted marginal effects

Table 5 presents the predicted marginal effects of each policy on the probabilities of choosing the lowest and highest categories of second-child intentions. For monetary incentives, each additional 100 CNY was associated with a 2.1 percentage point reduction in the probability of reporting “no intention of having a second child” (response category 0) (95% CI -2.2 to -1.9), and a 0.7 percentage point increase in the probability of choosing “absolutely want a second child” (response category 5) (95% CI 0.6 to 0.7).

**Table 5:** Marginal effects of choosing categories representing lowest and highest intentions to have a second child.

intentions to have a second child
| | **Lowest Category**<br>($DCE_{it} = 0$) | | **Highest Category**<br>($DCE_{it} = 5$) | |
| --- | --- | --- | --- | --- |
|  | Prob | 95% CI | Prob | 95% CI |
| ***Baby stipend (effect in 100s of CNY)*** | -.021*** | [-.022, -.019] | .007*** | [.006, .007] |
| ***Paid maternity leave*** |  |  |  |  |
| 98 days |  | *Reference Level* |  |  |
| 158 days | -.069*** | [-.080, -.058] | .018*** | [.015, .020] |
| 270 days | -.134*** | [-.148, -.120] | .045*** | [.039, .051] |
| ***Paid paternity leave*** |  |  |  |  |
| None |  | *Reference Level* |  |  |
| 20 days | -.100*** | [-.110, -.089] | .026*** | [.022, .030] |
| 98 days | -.126*** | [-.138, -.114] | .037*** | [.032, .042] |
| ***Childcare support*** |  |  |  |  |
| None |  | *Reference Level* |  |  |
| Free access to crèche 2 days a week for 3 years | -.050*** | [-.060, -.040] | .014*** | [.011, .017] |
| Free access to crèche 5 days a week for 3 years | -.092*** | [-.104, -.079] | .031*** | [.025, .036] |
| ***Career progression*** |  |  |  |  |
| Promotion delayed by 2 years |  | *Reference Level* |  |  |
| Promotion delayed by 1 year | -.048*** | [-.059, -.037] | .010*** | [.007, .012] |
| Promotion not affected | -.150*** | [-.164, -.137] | .050*** | [.044, .056] |

CI = Confidence Interval
\* $p < 0.05$ , \*\* $p < 0.01$ , \*\*\* $p < 0.001$

Career progression exerted the strongest effect. Preserving existing promotion opportunities (vs. a two-year delay) was associated with a 15.0 percentage point decrease (95% CI -16.4 to -13.7) in the probability of no intention and a 5.0 percentage point increase (95% CI 4.4 to 5.6) in definite intention.

Parental leave exhibited the second-largest impact. Extending maternity leave from 98 to 270 days reduced the probability of no intention by 13.4 percentage points (95% CI -14.8 to -12.0) and increased the probability of definite intention by 4.5 percentage points (95% CI 3.9 to 5.1). For paternity leave, extending from 0 to 20 days decreased the probability of no intention by 10.0 percentage points (95% CI -11.0 to -8.9) and increased the probability of definite intention by 2.6 percentage points (95% CI 2.3 to 3.0).

Providing free access to childcare five days per week was associated with a 9.2 percentage point decrease (95% CI -10.4 to -7.9) in the probability of no intention, and a 3.1 percentage point increase (95% CI 2.5 to 3.6) in the probability of the highest intention, compared to no access.

### 3.5 Heterogeneity and Sensitivity Analysis

Almost all estimates of the impact of changes in family support policy were robust to heterogeneity and sensitivity analyses, as shown in Appendices C-F. Particularly noteworthy is the fact that all policy attributes showed greater impact on respondent’s fertility preferences among higher response categories (such as “absolutely want another child”) than lower ones (such as “absolutely do not want another child”). We found evidence for interaction effects between policy attributes, but their magnitudes were much smaller than the main effects.

## 4 DISCUSSION

We conducted a single-profile DCE to elicit women’s preferences for family formation support policies in China. Overall, our findings reveal that women’s fertility decisions are highly sensitive to threats to their career progression, followed by variation in paid maternity and paternity leave, presence of childcare support, and provision of monetary incentives. These preferences remain robust even after accounting for heterogeneous effects and response inertia. Our results suggest that working environment reforms that balance women’s aspirations for career and family, and equalise gendered career opportunities, may be the most effective strategy to improve fertility intentions in China. Gender-egalitarian reforms in Nordic countries have demonstrated success in this regard (Rovny, 2011), and similar expectations have been observed in other low-fertility Asian settings (Gauthier and Gietel-Basten, 2024, Thévenon and Gauthier, 2011, Thévenon, 2011). While public childcare services are often regarded as effective in relieving women from caregiving responsibilities and enabling their career potential, our study indicates that their overall impact in China may be relatively limited. In addition, we found that women expect their spouses to take on a larger role in childbearing, highlighting the need for further research into how this expectation might influence men’s fertility intentions.

Career implications emerged as a central factor influencing married women’s decisions to have a second child. Among all attributes, respondents placed the highest value on the support of eliminating career penalties associated with childbearing, highlighting a strong aversion to career setbacks. This aligns with previous qualitative findings indicating that many women perceive achieving significant career milestones as a prerequisite to have a second child (Zhou, 2019). In contemporary China, while advancements in gender equality in job market and higher educational attainment among women have resulted in a societal expectation for women to participate in the workforce, traditional gender norms at the family level persist, placing a disproportionate burden of childcare and housework on working women (Jones, 2019). The pressure to excel in both domains make women prioritise career aspirations over the second child, with their first child already satisfying prenatal social norms after marriage. This dynamic reflects similar patterns observed in European countries during their respective demographic transitions, where greater gender equity in childcare responsibilities and career opportunities have subsequently led to fertility rebounds (Morgan and Taylor, 2006).

Our data also indicate a preference for more gender-equal set of family policy interventions. Women preferred extending paternity leave from 0 to 20 days over further extending maternity leave from 98 to 158 days, which may reflect a desire among married women for men to share childcare responsibilities during the initial postnatal period. Research in low-fertility countries, including both high-income and middle-income contexts, suggests that the sharing of childcare responsibilities is fast becoming a major driver of fertility intention among women (Sheppard, 2024, Séjourné et al., 2012). In addition, it may also reflect a desire among women for a more equitable working environment in China. Existing gendered parental leave durations – 98 days for women compared to none for men under legal entitlements – lead employers to favour male candidates to avoid the costs associated with extended maternity leave (Zhou, 2019). We found extending paternity by 20 days can decrease the probability of no intention for a second child by 10.0 percentage points - an effect comparable to that of a 172-day extension of maternity leave. This significant high valuation of paternity leave suggests that paternity leave can be a highly cost-effective policy lever to address low fertility rates in China.

Childcare support policies were estimated to have the least impact on fertility intention, with a WTP of 444 CNY [62 USD] per month for free full-time access. However, the average market price of daycare in China is significantly higher - 1,978 CNY [275 USD] per month on average, and over 5,500 CNY [765 USD] in metropolitan cities where work-family conflict is particularly pronounced (China Daily, 2024). This substantial gap between WTP and actual costs may partly explain why the national daycare enrolment rate remains below 8%, despite government efforts to promote nursery services through operational subsidies and childcare tax deduction (China Daily, 2024). These findings further suggest that direct cash transfers to families might be a more cost-effective policy lever than offering subsidised or even free daycare services regarding improving fertility intentions. While government spending on daycare has markedly increased fertility rates in OECD countries (Rovny, 2011), the same success may not be replicable in China. The underlying reasons why women perceive childcare burdens as major barriers to fertility, but place relatively low value on free childcare services warrant further investigation.

Monetary incentives demonstrated a measurable, though low, effect on fertility intentions. An additional 100 CNY per month increased the probability of having a second child by 0.7 percentage points, equivalent to a 7% absolute increase with a monthly payment of 1,000 CNY [139 USD] over three years. This projection is slightly lower than the 8.5% increase estimated by a similar study in eastern China, which assessed broader fertility intentions, i.e. the second, third or more children (Dong et al., 2024). As we observed earlier, these studies focus on the *direct* costs of childbearing, rather than the *indirect* or *opportunity* costs which are inevitably much higher for females (and their career progression) in the context of an ‘incomplete gender revolution’ of unequal sharing of caring and domestic responsibilities (Esping-Andersen, 2009, McDonald, 2000, McDonald, 2013).

While all proposed policies showed significant impact on the fertility intention of having a second child, the magnitudes of impact in most cases remain small. The largest marginal gain in probability of expressing a definite desire for a second child was 5 percentage points, from ensuring no delay in one’s career progression, while the reductions in the probability of indicating no intention whatsoever of having a second child were larger (>10 percentage points). This indicates that, while these policies are perceived as helpful enough in making women *consider* having a second child, they may be insufficient in themselves to make a large number of married women actually *decide* to have a second child. Prior research suggests that family welfare policies tend to have stronger effects in high-fertility contexts than in low-fertility ones (Zhang et al., 2023). With China’s fertility rate persistently below the lowest-low threshold, broader structural reforms emphasising gender equity and workplace flexibility may be urgently needed.

### 4.1 Strengths and limitations

While the sample is geographically representative of the different provinces of China, it may not be fully representative across other dimensions, such as education level or urban/rural distribution. Specifically, the sample consists of a slightly more educated group with higher income, which may not fully reflect the national population. However, it is worth noting that highly educated women in China tend to have lower fertility intentions, making this subgroup particularly relevant for understanding the drivers and barriers to fertility behaviours (Xiang et al., 2023). Besides, there is a risk of self-selection bias, as participants were recruited through an online survey. But considering a response rate of 83.9%, the impact is expected to be limited. Finally, stated preferences would not necessarily translate to realised fertility behaviours (Philipov et al., 2009). This has been observed in various other DCE studies. Despite this, it has been widely demonstrated that preferences are a key part of the fertility decision-making process, and any exploration of their formation and the removal of barriers to their actualisation is still important (Philipov et al., 2009, Ajzen and Klobas, 2013, Ajzen, 1991), especially in the current policy context of China.

## 5. CONCLUSION

China is undoubtedly facing demographic headwinds over the next century. Urgent reform is needed in a variety of areas which will be squeezed by the twin processes of population ageing and decline. It is not unexpected that the government has focussed on tackling low fertility. Evidence from elsewhere, as well as recent evidence from China itself, suggests, however, a top-down search for a ‘demographic solution to a demographic problem’ may be ineffective. Despite this, efforts to reducing the barriers to meeting the reproductive aspirations for individuals can bring about a double benefit: enabling individuals to fulfil their reproductive goals while enhancing other aspects of work and life; potentially slowing the pace and scale of population ageing and decline.

Our results highlight the critical role of career progression in shaping fertility decisions and thus the need for an emphasis on gender-responsive policy interventions. Although overall marginal effects remained modest, suggesting only small shifts are possible under existing policy proposals, our findings point towards policies promoting gender equity at work, such as protection for women’s career progression and gender-equal parental leave packages may be the most promising avenues for removing barriers for women (and their partners) to meet their reproductive aspirations. These findings offer key policy levers for Chinese policymakers who are concerned about future demographic changes to align with women’s preferences and support their family formation.

## Data Availability

All data produced in the present study are available upon reasonable request to the authors

## Acknowledgements

We thank the UCL-PKU Strategic Fund for financial support with this project.

## Footnotes

1 https://www.fujian.gov.cn/english/news/202108/t20210809_5665713.htm

2 https://clb.org.hk/en/content/challenges-and-concerns-surrounding-chinas-retirement-age-reform

3 https://www.globaltimes.cn/page/202405/1312184.shtml

4 CNY was converted to U.S. dollars (USD) according to the average exchange rate in 2022, which was 1 USD = 7.189 CNY. Data found at https://www.irs.gov/individuals/international-taxpayers/yearly-average-currency-exchange-rates

## References

Ajzen, I. 1991. The Theory of planned behavior. Organizational Behavior and Human Decision Processes.

Ajzen, I. & Klobas, J. 2013. Fertility intentions: An approach based on the theory of planned behavior. Demographic research, 29, 203–232.

Bloom, D. E., Canning, D. & Lubet, A. 2015. Global Population Aging: Facts, Challenges, Solutions & Perspectives. Daedalus, 144, 80–92.

Bridges, J. F., Hauber, A. B., Marshall, D., Lloyd, A., Prosser, L. A., Regier, D. A., Johnson, F. R. & Mauskopf, J. 2011. Conjoint analysis applications in health—a checklist: a report of the Ispor Good Research Practices for Conjoint Analysis Task Force. Value in health, 14, 403–413.

Caldwell, J. C. & Caldwell, B. K. 2005. The Causes Of The Asian Fertility Decline. Asian Population Studies, 1, 31–46.

Chen, Q., Wang, A., Song, X., Liu, X., Liu, Y., Wei, J., Shu, J., Sun, M., Zhong, T. & Luo, M. 2023. Fertility intentions to have a second or third child among the childbearing-age population in Central China under China’s three-child policy: a cross-sectional study. Journal of Global Health, 13.

Chen, S. & Gietel-Basten, S. 2024. How genuine are sub-replacement ideal family sizes in urban China? Population Studies, 78.

China Daily. 2024. More childcare services nationwide aim to reduce burden on families [Online]. Available: https://www.chinadaily.com.cn/a/202409/13/WS66e39be1a3103711928a7d09.html [Accessed 13 February 2025].

Choi, S.-W., Yellow Horse, A. J. & Yang, T.-C. 2018. Family policies and working women’s fertility intentions in South Korea. Asian Population Studies, 14, 251–270.

Clark, M. D., Determann, D., Petrou, S., Moro, D. & De Bekker-Grob, E. W. 2014. Discrete choice experiments in health economics: a review of the literature. Pharmacoeconomics, 32, 883–902.

Coleman, D. & Basten, S. 2015. The Death of the West: An alternative view. Population Studies, 69, S107–S118.

De Bekker-Grob, E. W., Donkers, B., Jonker, M. F. & Stolk, E. A. 2015. Sample Size Requirements for Discrete-Choice Experiments in Healthcare: a Practical Guide. The Patient-Patient-Centered Outcomes Research, 8, 373–384.

Dell’olio, L., Ibeas, A., Oña, J. D. & Oña, R. D. 2018. Methods Based on Random Utility Theory. Elsevier.

Díaz LUévano, C., Sicsic, J., Pellissier, G., Chyderiotis, S., Arwidson, P., Olivier, C., Gagneux-Brunon, A., Botelho-Nevers, E., Bouvet, E. & Mueller, J. 2021. Quantifying healthcare and welfare sector workers’ preferences around Covid-19 vaccination: a cross-sectional, single-profile discrete-choice experiment in France. Bmj Open, 11, e055148.

Doepke, M., Hannusch, A., Kindermann, F. & Tertilt, M. 2023. The economics of fertility: A new era. Handbook of the Economics of the Family. Elsevier.

Dong, W.-H., Wang, X., Yuan, F., Wang, L., Gu, T.-M., Zhu, B.-Q. & Shao, J. 2024. Will a government subsidy increase couples’ further fertility intentions? A real-world study from a large-scale online survey in Eastern China. Human Reproduction Open, 2024.

Dorling, D. & Gietel-Basten, S. 2017. Why demography matters, John Wiley & Sons.

Esping-Andersen, G.2009. Incomplete revolution: Adapting welfare states to women’s new roles, Polity.

Gauthier, A. & Gietel-Basten, S. 2024. Family Policy and Fertility: Evidence and Reflections[forthcoming]. Population and Development Review.

Gietel-Basten, S. 2023. Population aging in China from a multidimensional, comparative perspective. China Population and Development Studies, 7, 104–110.

Gietel-Basten, S., Han, X. & Cheng, Y. 2019. Assessing the impact of the “one-child policy” in China: a synthetic control approach. PLos one, 14, e0220170.

Gietel-Basten, S., Rotkirch, A. & Sobotka, T. 2022. Changing the perspective on low birth rates: why simplistic solutions won’t work. bmj, 379.

Goodkind, D. 2017. The astonishing population averted by China’s birth restrictions: Estimates, nightmares, and reprogrammed ambitions. Demography, 54, 1375–1400.

Janssen, E. M., Hauber, A. B. & Bridges, J. F. 2018. Conducting a discrete-choice experiment study following recommendations for good research practices: an application for eliciting patient preferences for diabetes treatments. Value in Health, 21, 59–68.

Jones, G., Straughan, P. T. & Chan, A. 2009. Ultra-low fertility in Pacific Asia. Trends, Causes and Policy lssues, *Abingdon, Oxon, Uk*: *Routledge*.

Jones, G. W. 2019. Ultra-low fertility in East Asia: policy responses and challenges. Asian Population Studies, 15, 131–149.

Li, Y. & Jiang, Q. 2019. Women’s gender role attitudes and fertility intentions of having a second child: survey findings from Shaanxi Province of China. Asian Population Studies, 15, 66–86.

Liu, J., Liu, M., Zhang, S., Ma, Q. & Wang, Q. 2020. Intent to have a second child among Chinese women of childbearing age following China’s new universal two-child policy: a cross-sectional study. Bmj sexual & reproductive health, 46, 59–66.

Lopreite, M. & Zhu, Z. 2020. The effects of ageing population on health expenditure and economic growth in China: A Bayesian-Var approach. Social science & medicine, 265, 113513.

Mcdonald, P. 2000. Gender equity, social institutions and the future of fertility. Journal of the Australian Population Association, 17, 1–16.

Mcdonald, P. 2013. Societal foundations for explaining low fertility: Gender equity. Demographic research, 28, 981–994.

Mitter, R. 2024. Open or closed? China’s dilemmas in a changing geopolitical and geoeconomic order. Oxford Review of Economic Policy, 40, 366–373.

Morgan, S. P. & Taylor, M. G. 2006. Low Fertility at the Turn of the Twenty-First Century. Annual Review of Sociology, 32, 375–399.

Peng, I. & Yeandle, S. 2017. Eldercare policies in East Asia and Europe: Mapping policy changes and variations and their implications.

Peng, X. 2021. Coping with population ageing in mainland China. Asian Population Studies, 17, 1–6.

Philipov, D., THévenon, O., Klobas, J., Bernardi, L. & Liefbroer, A. C. 2009. Reproductive decision-making in a macro-micro perspective (Repro): State-of-the-art review. European Demographic Research Papers, 1.

Rovny, A. E. 2011. Welfare state policy determinants of fertility level: A comparative analysis. Journal of European Social Policy, 21, 335–347.

SéJOURNé, N., Vaslot, V., BEAUMé, M., Goutaudier, N. & Chabrol, H. 2012. The impact of paternity leave and paternal involvement in child care on maternal postpartum depression. Journal of Reproductive and Infant Psychology, 30, 135–144.

Shen, K., Wang, F. & Cai, Y. 2020. Government policy and global fertility change: a reappraisal. Asian Population Studies, 16, 145–166.

Sheppard, P. 2024. Using discrete choice modeling to understand the drivers of reproductive delay in the United Kingdom. International Journal of Population Studies, 0, 3600.

Stowe, R. 2005. Aging China: The demographic challenge to China’s economic prospects, Bloomsbury Publishing Usa.

THévenon, O. 2011. Family policies in Oecd countries: A comparative analysis. Population and development review, 37, 57–87.

THévenon, O. & Gauthier, A. H. 2011. Family policies in developed countries: A ‘fertility-booster’with side-effects. Community, Work & Family, 14, 197–216.

Undesa. 2024. 2024 Revision of World Population Prospects [Online]. Available: https://population.un.org/wpp/ [Accessed 13 February 2025].

Wang, F., Cai, Y., Shen, K. & Gietel-Basten, S. 2018. Is demography just a numerical exercise? Numbers, politics, and legacies of China’s one-child policy. Demography, 55, 693–719.

Wang, S., Ren, Z., Xiao, Z., Wang, N., Yang, H. & Pu, H. 2022. Coupling analysis of population aging and economic growth with spatial-temporal variation: a case study in China. International Journal for Equity in Health, 21, 107.

Wang, Y., Kong, F., Fu, Y. & Qiao, J. 2024. How can China tackle its declining fertility rate? bmj, 386.

West, R. & Ramcharan, P. 2019. The emerging role of Financial Counsellors in supporting Older Persons in financial hardship and with management of Consumer - directed Care packages within Australia. Australian Journal of Social Issues, 54, 32–51.

Williams, R. 2016. Understanding and interpreting generalized ordered logit models. The Journal of Mathematical Sociology, 40, 7–20.

Xiang, Z., Zhang, X., Li, Y., Li, J., Wang, Y., Wang, Y., Ming, W.-K., Sun, X., Jiang, B. & Zhai, G. 2023. Fertility intention and its affecting factors in China: a national cross-sectional survey. Heliyon, 9.

Yang, Y., He, R., Zhang, N. & Li, L. 2023. Second-child fertility intentions among urban women in China: a systematic review and meta-analysis. International Journal of Environmental Research and Public Health, 20, 3744.

Zhai, Z., Jin, G. & Zhenwu Zhai, G. J. 2023. China’s family planning policy and fertility transition. Chinese Journal of Sociology, 9.

Zhang, T.-T., Cai, X.-Y., Shi, X.-H., Zhu, W., Shan, S.-N., Zhang, T.-T., Cai, X.-Y., Shi, X.-H., Zhu, W. & Shan, S.-N. 2023. The Effect of Family Fertility Support Policies on Fertility, Their Contribution, and Policy Pathways to Fertility Improvement in OECD Countries. International Journal of Environmental Research and Public Health 2023, *Vol*. 20, *Page* 4790, 20.

Zhang, X. & Chand, S. 2024. An Analysis of the Chinese Pension System Pointing to the Urgent Need for Reform. The Chinese Economy, 1–15.

Zhou, X., Li, Y., Correa, A., Salustri, F. & Skordis, J. 2023. The need for voices from the grassroots in China’s public health system. The Lancet Regional Health – Western Pacific, 32.

Zhou, Y. 2019. The dual demands: Gender equity and fertility intentions after the one-child policy. Journal of Contemporary China, 28, 367–384.

