## Appendix for "What Family Policies do Chinese Women Want? A Discrete Choice Experiment"

**Supplementary Appendices**

**Appendix A: Justification for DCE Attribute Levels**

We followed the International Society for Pharmacoeconomics and Outcomes Research (ISPOR) Conjoint Analysis Task Force guidelines for experimental design and selection of attributes and levels (Bridges et al., 2011).

***Maternity Leave (3 Levels)***

We used the current legal entitlement of 98 days in China as the reference level, with additional levels set at 158 days (piloted in some provinces in China) and 270 days (the third-highest level globally) (World Population Review, 2024). This range was selected to ensure sufficient variation for estimating the impact of maternity leave on women’s decisions and preferences. While longer leave is beneficial to maternal and child health (Staehelin et al., 2007), it can also cause significant work interruptions that can affect career advancement (Binaku-Hajrullahu, 2019).

***Paternity Leave (3 Levels)***

The reference level for paternity leave was the status quo, which is minimal or non-existent in most cases. We added levels consisting of 20 days’ leave (piloted in some provinces) and 98 days’ leave (equal to current statutory maternity leave).

***Childcare Support (3 Levels)***

This attribute described the number of days per week that free crèche services could be made available for children aged 0-3. Since lack of childcare is often cited as a leading reason deterring women from having more children (Jing et al., 2022), we used this attribute to explore whether public childcare services could positively influence fertility intentions. Daycare support in countries like Japan, South Korea and Singapore have demonstrated some impact, but it remains uncertain what *level* of support might shift women’s fertility intentions (Choi et al., 2018; Jones, 2019). To gauge respondents' sensitivity to this policy, we included three levels: no free crèche, two days per week, and five days per week (aligned with standard working days) for the first three years of the child’s life.

***Monetary Incentives (4 Levels)***

This attribute was designed to quantify how financial incentives might influence fertility intentions. We framed it as a ‘childcare subsidy’ and specified levels at 0 CNY, 500 CNY, 1000 CNY, and 1500 CNY per month per child under age three. The amounts were selected with reference to Korea’s childcare subsidies, scaled in proportion to China’s GDP per capita (Li et al., 2024), and rounded to the nearest multiple of 500 to simplify participant evaluations.

***Career Impact (3 Levels)***

This attribute tested the extent to which potential career impact of childbearing would influence women’s fertility intentions. We specified the following levels: no impact on career progression, promotion delayed by one year, and promotion delayed by two years. While career progression is subject to various real-world factors and may beyond the reach of policy interventions, we used this attribute as context to understand whether lowering the career opportunity cost of childbearing could significantly affect women’s intention to have more children.

**Appendix B: Proportional odds model for constant preference weights**

Recall that we modelled respondents’ utility using the equation $U_{i}=\sum_{\boldsymbol{j=1}}^{\boldsymbol{5}} \beta_{j}X_{ij}+u_{i}+v_{i}, u_{i}\sim N(0, \sigma_{u}^{2})$ (see section 2.4). The fertility choice was modelled using the equation:

$$\begin{aligned} logit\left（ \Pr\left( DCE_{i}>m | X_{ij} \right) \right）=logit\left（ Pr\left( U_{i}>\kappa_{m-1} | X_{ij} \right) \right）=\sum_{\boldsymbol{j=1}}^{\boldsymbol{5}} \beta_{j}X_{ij}+u_{i}-\kappa_{m-1}\#\left( 1 \right) \end{aligned}$$

where $logit(p)=ln(\frac{p}{1-p})$, $X_{ij}$ is one of the five policy attributes and$\beta_{j}$ is the corresponding preference weight. In this model, which we used for the main analysis, the effect of policy attributes, $X_{ij},$ on the change in log-odds is assumed to be constant across all cutoff points. Further, $DCE_{i}$ represents the fertility intention of the respondent using response categories ranging from $m = 0$ ("no intention of having a second child") to $m = 5$ ("definitely want a second child"). $\kappa_{m}$ for $m=1,2,3,4,5$are cutoff points for response categories, translating greater utility $U_{i}$ into higher response categories $m.$ In particular, the probability of a respondent’s fertility intention falling above category $m$ equals the probability of their utility $U_{i}$exceeding the corresponding cutoff point $\kappa_{m-1}$, given the family policy packages. Finally, $u_{i}$ is an individual-level random effect and $v_{i}$ is an error term.

**Appendix C: Partial proportional odds model for heterogeneous preference weights**

We relaxed the assumption of constant changes in the log-odds of intending to have another child using a partial proportional odds model instead of a canonical proportional odds model. We now modelled fertility intention using the following equation

$$\begin{aligned} logit (\Pr\left( DCE_{i}>m | \boldsymbol{Z}_{\boldsymbol{i}}\boldsymbol{,}\boldsymbol{W}_{\boldsymbol{i}} \right))=logit (Pr \left( U_{i}>\kappa_{m-1}|\boldsymbol{Z}_{\boldsymbol{i}}\boldsymbol{,}\boldsymbol{W}_{\boldsymbol{i}} \right))=\boldsymbol{Z}_{\boldsymbol{i}}\boldsymbol{\beta+}\boldsymbol{W}_{\boldsymbol{i}}\boldsymbol{\gamma}_{m-1}-\kappa_{m-1}\#\left( 2 \right) \end{aligned}$$

Here, most of the variables ($DCE_{i}, m, U_{i}, \kappa_{m-1}$) have the same meaning as in equation (1). However, the five family policies $X_{ij}$ are now separated into policy attributes with constant impact across all response categories (represented by vector $\boldsymbol{Z}_{\boldsymbol{i}}$) and policy attributes with varying impacts across response categories (represented by a vector $\boldsymbol{W}_{\boldsymbol{i}}$). We compared a fully constrained model (all policy attributes in $\boldsymbol{Z}_{\boldsymbol{i}}$) with a fully unconstrained model (all policy attributes in $\boldsymbol{W}_{\boldsymbol{i}}$) and chose an intermediary model through backwards elimination using gologit2, autofit in Stata.

Using this model, we were able to explore whether policies had differential impacts on women with low versus high fertility intention at baseline. The results, reported in Table A1, can be read as follows. First, the baby stipend, having free access to a crèche 2 days a week for 3 years, and having a promotion delayed by 1 year all exerted constant impact on fertility intentions in the final model, as shown in first column. Second, for policies with differential impacts, the right five columns show the impact estimates for each cutoff.

For instance, we found that extending maternity leave from 98 to 270 days made respondents 5.82 times more likely to report “definitely want a second child” (response category 5) than any of the other levels of fertility intention (response categories 0-4). However, the same extension only led to a 2.30 times reduction in the likelihood of reporting “no intention of having a second child” (response category 0) relative to any of the other levels of fertility intention (categories 1-5). This shows that extending maternity leave from 98 to 270 days was more effective at encouraging high levels of fertility intention than discouraging low levels of fertility intention.

| **Table A1. Estimated preference weights using a partial proportional odds model** | | | | | | | | | | | | |
| --- | --- | --- | --- | --- | --- | --- | --- | --- | --- | --- | --- | --- |
|  | **CONSTANT impact** | | **VARYING impact** | | | | | | | | | |
|  | **across all cutoffs** | | **(1,2,3,4, and 5) vs 0** | | **(2, 3, 4, and 5) vs (0 and 1)** | | **(3,4, and 5) vs (0, 1, and 2)** | | **(4 and 5) vs (0, 1, 2 and 3)** | | **5 vs (0, 1, 2, 3 and 4)** | |
|  | OR | 95% CI | OR | 95% CI | OR | 95% CI | OR | 95% CI | OR | 95% CI | OR | 95% CI |
| ***Baby stipend (effect in 100s of CNY)*** | 1.12*** | [1.11, 1.14] |  |  |  |  |  |  |  |  |  |  |
| ***Paid maternity leave*** |  |  |  |  |  |  |  |  |  |  |  |  |
| 98 days | *Reference Level* | | | | | | | | | | | |
| 158 days |  |  | 1.49^***^ | [1.34,1.65] | 1.56^***^ | [1.42,1.71] | 1.72^***^ | [1.55,1.91] | 1.85^***^ | [1.60,2.14] | 2.23^***^ | [1.66,2.99] |
| 270 days |  |  | 2.30^***^ | [1.96,2.71] | 2.46^***^ | [2.14,2.82] | 2.93^***^ | [2.51,3.43] | 3.76^***^ | [3.06,4.61] | 5.82^***^ | [4.11,8.24] |
| ***Paid paternity leave*** |  |  |  |  |  |  |  |  |  |  |  |  |
| None | *Reference Level* | | | | | | | | | | | |
| 20 days |  |  | 1.62^***^ | [1.47,1.78] | 1.72^***^ | [1.59,1.86] | 1.75^***^ | [1.59,1.93] | 2.07^***^ | [1.79,2.38] | 2.36^***^ | [1.75,3.18] |
| 98 days |  |  | 1.91^***^ | [1.72,2.13] | 1.94^***^ | [1.77,2.11] | 2.21^***^ | [1.99,2.46] | 2.77^***^ | [2.40,3.20] | 3.10^***^ | [2.41,3.98] |
| ***Childcare support*** |  |  |  |  |  |  |  |  |  |  |  |  |
| None | *Reference Level* | | | | | | | | | | | |
| Free access to crèche 2 days a week for 3 years | 1.48*** | [1.38, 1.59] |  |  |  |  |  |  |  |  |  |  |
| Free access to crèche 5 days a week for 3 years |  |  | 1.88^***^ | [1.65,2.13] | 1.70^***^ | [1.53,1.89] | 1.78^***^ | [1.58,2.00] | 2.02^***^ | [1.75,2.34] | 2.75^***^ | [2.18,3.48] |
| ***Career progression*** |  |  |  |  |  |  |  |  |  |  |  |  |
| Promotion delayed by 2 years | *Reference Level* | | | | | | | | | | |  |
| Promotion delayed by 1 year | 1.62*** | [1.47, 1.79] |  |  |  |  |  |  |  |  |  |  |
| Promotion not affected |  |  | 1.93^***^ | [1.68,2.22] | 2.44^***^ | [2.17,2.74] | 3.39^***^ | [3.00,3.83] | 3.63^***^ | [3.11,4.24] | 4.35^***^ | [3.33,5.67] |
| OR = Odds Ratio, CI = Confidence Interval  ^*^ *p* < 0.05, ^**^ *p* < 0.01, ^***^ *p* < 0.001  0 = absolutely don’t want a second child, gradually progresses through categories 1, 2, 3 and 4, to 5 = absolutely want a second child. | | | | | | | | | | | | |

**Appendix D: Sensitivity Analyses Accounting for Decision Inertia**

To analyse our model’s sensitivity to decision inertia (a phenomenon in which choices made in previous DCE scenarios influence subsequent choices in subsequent scenarios), we used two methods. First, we included responses from the immediately prior choice task as a lagged dependent variable. We fitted the following model:

$$\begin{aligned} logit\left（ \Pr\left( DCE_{i}>m | X_{ij} \right) \right）=logit\left（ Pr\left( U_{i}>\kappa_{m-1} | X_{ij} \right) \right）=\sum_{\boldsymbol{j=1}}^{\boldsymbol{5}} \beta_{j}X_{ij}+DC{E^{'}}_{i}+u_{i}-\kappa_{m-1} \#\left( 3 \right) \end{aligned}$$

where most variables have the same meaning as in equation (1), but we added a term $DC{E'}_{i}$ , representing the respondent $i$’s choice in the previous DCE task. Results are reported in Table A2 – Column B.

Second, we excluded respondents who consistently expressed low desirability for a second child (response categories 0-2) across all 12 scenarios or alternatively expressed high desirability (response categories 3-5) across all 12 scenarios and re-estimated equation (1) using the resultant subsample. Results are reported in Table A2 – Column C.

| **Table A2: Sensitivity Analysis Using Models Incorporating Decision Inertia** | | | |
| --- | --- | --- | --- |
|  | Column A | Column B | Column C |
|  | Base Model -  Equation (1) | Model with a lagged response - Equation (3) | Model excluding disengaged responses |
| ***Baby stipend (effect in 100s of CNY)*** | 1.23^***^ | 1.24^***^ | 1.25^***^ |
|  | [1.22,1.25] | [1.22,1.27] | [1.23,1.27] |
| ***Paid maternity leave*** | *Reference Level* | | |
| 98 days |  |  |  |
| 158 days | 1.92^***^ | 1.84^***^ | 2.01^***^ |
|  | [1.74,2.12] | [1.66,2.04] | [1.80,2.24] |
| 270 days | 3.96^***^ | 4.03^***^ | 4.33^***^ |
|  | [3.44,4.56] | [3.49,4.65] | [3.70,5.06] |
| ***Paid paternity leave*** | *Reference Level* | | |
| None |  |  |  |
| 20 days | 2.60^***^ | 2.54^***^ | 2.88^***^ |
|  | [2.35,2.87] | [2.28,2.82] | [2.59,3.20] |
| 98 days | 3.49^***^ | 3.37^***^ | 3.99^***^ |
|  | [3.11,3.91] | [3.01,3.78] | [3.50,4.55] |
| ***Childcare support*** | *Reference Level* | | |
| None |  |  |  |
| Free access to crèche 2 days a week for 3 years | 1.62^***^ | 1.59^***^ | 1.70^***^ |
|  | [1.48,1.78] | [1.44,1.77] | [1.54,1.89] |
| Free access to crèche 5 days a week for 3 years | 2.55^***^ | 2.52^***^ | 2.66^***^ |
|  | [2.24,2.90] | [2.21,2.87] | [2.29,3.08] |
| ***Career progression*** | *Reference Level* | | |
| Promotion delayed by 2 years |  |  |  |
| Promotion delayed by 1 year | 1.55^***^ | 1.55^***^ | 1.61^***^ |
|  | [1.40,1.70] | [1.39,1.72] | [1.44,1.79] |
| Promotion not affected | 4.76^***^ | 4.79^***^ | 5.67^***^ |
|  | [4.16,5.45] | [4.17,5.50] | [4.87,6.60] |
| Lagged DCE response |  | 1.09^***^ |  |
|  |  | [1.05,1.14] |  |
| ***N*** | 1005 | 921 | 742 |
| Reported estimates are odds ratio, with 95% confidence interval in the squared bracket below.  ^*^ *p* < 0.05, ^**^ *p* < 0.01, ^***^ *p* < 0.001 | | | |

**Appendix E: Interaction Effects Between Policy Attributes and Individuals’ Characteristics**

To explore whether different family policies might interact when producing impacts on women's fertility intention, we estimated an expanded model incorporating interaction effects. We used the following specification:

$$\begin{aligned} logit\left（ \Pr\left( DCE_{i}>m | X_{ij} \right) \right）=logit\left（ Pr\left( U_{i}>\kappa_{m-1} | X_{ij} \right) \right） \\ =\sum_{\boldsymbol{j=1}}^{\boldsymbol{5}} \beta_{j}X_{ij}+ \sum_{k=1}^{4} \sum_{\boldsymbol{l=k+1}}^{\boldsymbol{5}} \omega_{kl}{(X}_{ik}\times X_{il})+u_{i}-\kappa_{m-1}\#\left( 4 \right) \end{aligned}$$

where most of the notation follows equation (1), but we added terms $X_{ik}\times X_{il}$ representing **pairwise combinations** of the five family policies and $\omega_{kl}$ denoting the size of the interaction effect. The results are summarised in Table A3 - Column B.

To investigate whether the impact of policy changes on fertility intentions further varied according to women’s individual characteristics, we estimated an additional model that includes interaction terms between policy attributes and factors such as annual household income, ideal number of children, feasible number of children, and intergenerational childcare support. The model used the following specification:

$$\begin{aligned} logit\left（ \Pr\left( DCE_{i}>m | X_{ij} \right) \right）=logit\left（ Pr\left( U_{i}>\kappa_{m-1} | X_{ij} \right) \right） \\ =\sum_{\boldsymbol{j=1}}^{\boldsymbol{5}} \beta_{j}X_{ij}+\sum_{\boldsymbol{d=1}}^{\boldsymbol{4}} \theta_{d}Q_{id}+\sum_{j=1}^{5} \sum_{\boldsymbol{d=1}}^{\boldsymbol{4}} \sigma_{kl}{(X}_{ij}\times Q_{id})+u_{i}-\kappa_{m-1} \\ \#\left( 5 \right) \end{aligned}$$

where most of the notation follows equation (1), but we added covariates $Q_{id}$, representing respondent $i$’s personal characteristics and $\sigma_{kl}$ representing the size of the interaction effects between each policy attribute and each personal characteristic.

Model results are reported in Table A3 - Column C. While interaction between policy attributes were statistically significant, their magnitudes were much smaller compared to main effects. Notably, simultaneous extensions of maternity leave to 270 days and paternity leave to 98 days were associated with an additional 1.69 times higher odds (P < 0.05, 95% CI: 1.03–2.76) of leaning towards having a second child. Change in annual household income did not show a measurable modification effect in the relationships between policy attributes and fertility intentions. Considering two children instead of one as ideal or feasible amplified the impact of extended paternity leave, but did not modify other policies’ effect. As expected, currently having grandparental childcare support significantly reduced the effect of public crèche services in increasing fertility intentions. The estimation of the impacts of all families polices stayed robust after the inclusion of all these interaction terms.

| **Table A3: Interaction Effects** **Between Policy Attributes and** **Individuals’ Characteristics** | | | | | | |
| --- | --- | --- | --- | --- | --- | --- |
|  | Column A | | | Column B | Column C | |
|  | Base Model -  Equation (1) | | | Interactional policy Attributes | Interactional Attitudes | |
| ***Baby stipend (effect in 100s of CNY)*** | 1.23^***^ | | | 1.37^***^ | 1.22^***^ | |
|  | [1.22,1.25] | | | [1.26,1.48] | [1.17,1.27] | |
| ***Paid maternity leave*** | *Reference Level* | | | | | |
| 98 days |  |  |  |  |  |  |
| 158 days | 1.92^***^ | | | 3.24^***^ | 1.96^***^ | |
|  | [1.74,2.12] | | | [2.10,4.99] | [1.45,2.66] | |
| 270 days | 3.96^***^ | | | 6.04^***^ | 3.18^***^ | |
|  | [3.44,4.56] | | | [3.44,10.60] | [1.96,5.17] | |
| ***Paid paternity leave*** | *Reference Level* | | | | | |
| None |  |  |  |  |  |  |
| 20 days | 2.60^***^ | | | 4.71^***^ | 1.52^*^ | |
|  | [2.35,2.87] | | | [2.93,7.60] | [1.04,2.20] | |
| 98 days | 3.49^***^ | | | 9.37^***^ | 2.08^***^ | |
|  | [3.11,3.91] | | | [5.73,15.31] | [1.41,3.06] | |
| ***Childcare support*** | *Reference Level* | | | | | |
| None |  |  |  |  |  |  |
| Free access to crèche 2 days a week for 3 years | 1.62^***^ | | | 1.16 | 1.60^**^ | |
|  | [1.48,1.78] | | | [0.80,1.66] | [1.18,2.17] | |
| Free access to crèche 5 days a week for 3 years | 2.55^***^ | | | 0.57 | 3.89^***^ | |
|  | [2.24,2.90] | | | [0.28,1.16] | [2.39,6.31] | |
| ***Career progression*** | *Reference Level* | | | | | |
| Promotion delayed by 2 years |  |  |  |  |  |  |
| Promotion delayed by 1 year | 1.55^***^ | | | 1.32 | 1.44^*^ | |
|  | [1.40,1.70] | | | [0.97,1.81] | [1.04,1.99] | |
| Promotion not affected | 4.76^***^ | | | 11.88^***^ | 2.79^***^ | |
|  | [4.16,5.45] | | | [7.10,19.89] | [1.91,4.07] | |
| ***Interactional effects between policies*** |  | | |  |  | |
| Baby stipend # Maternity leave of 270 days |  | | | 0.87^***^ |  | |
|  |  | | | [0.81,0.94] |  | |
| Baby stipend # Paternity leave of 98 days |  | | | 0.92^***^ |  | |
|  |  | | | [0.87,0.96] |  | |
| Baby stipend # Crèche 2 days a week |  | | | 1.06^*^ |  | |
|  |  | | | [1.00,1.11] |  | |
| Baby stipend # Crèche 5 days a week |  | | | 1.26^**^ |  | |
|  |  | | | [1.09,1.46] |  | |
| Baby stipend # Promotion not affected |  | | | 0.84^***^ |  | |
|  |  | | | [0.78,0.90] |  | |
| Maternity leave of 158 days # Paternity leave of 98 days |  | | | 0.60^**^ |  | |
|  |  | | | [0.43,0.85] |  | |
| Maternity leave of 270 days # Paternity leave of 98 days |  | | | 1.69^*^ |  | |
|  |  | | | [1.03,2.76] |  | |
| Maternity leave of 270 days # Crèche 2 days a week |  | | | 1.77^***^ |  | |
|  |  | | | [1.31,2.39] |  | |
| Crèche 5 days a week # Promotion delayed by 1 year |  | | | 2.26^**^ |  | |
|  |  | | | [1.30,3.93] |  | |
| Crèche 5 days a week # Promotion not affected |  | | | 1.70^*^ |  | |
|  |  | | | [1.02,2.83] |  | |
| ***Annual household income (effect in 10000s of CNY)*** | |  |  | | | 1.01 |
|  | |  |  | | | [1.00,1.01] |
| ***Grandparents’ childcare support*** | |  |  | | |  |
| Nearly none | |  |  | | | *Reference Level* |
| When necessary | |  |  | | | 0.71 |
|  | |  |  | | | [0.30,1.69] |
| Sometimes | |  |  | | | 1.20 |
|  | |  |  | | | [0.54,2.63] |
| Working days | |  |  | | | 1.13 |
|  | |  |  | | | [0.53,2.39] |
| Everyday | |  |  | | | 0.97 |
|  | |  |  | | | [0.46,2.05] |
| ***Feasible number of children*** | |  |  | | |  |
| 1 | |  |  | | | *Reference Level* |
| 0 | |  |  | | | 2.53 |
|  | |  |  | | | [0.05,140.95] |
| 2 | |  |  | | | 4.44^***^ |
|  | |  |  | | | [2.61,7.54] |
| More than 3 | |  |  | | | 5.54^*^ |
|  | |  |  | | | [1.35,22.70] |
| ***Ideal number of children*** | |  |  | | |  |
| 1 | |  |  | | | *Reference Level* |
| 0 | |  |  | | | 3.54 |
|  | |  |  | | | [0.05,270.36] |
| 2 | |  |  | | | 1.53 |
|  | |  |  | | | [0.86,2.74] |
| More than 3 | |  |  | | | 2.38 |
|  | |  |  | | | [0.92,6.15] |
| ***Interaction effects with household income*** | |  |  | | |  |
| Baby stipend # Annual household income | |  |  | | | 1.00^**^ |
|  | |  |  | | | [1.00,1.00] |
| Crèche 2 days a week # Annual household income | |  |  | | | 1.00^*^ |
|  | |  |  | | | [0.99,1.00] |
| Crèche 5 days a week # Annual household income | |  |  | | | 1.00^*^ |
|  | |  |  | | | [0.99,1.00] |
| ***Interaction effects with grandparents’ childcare support*** | |  |  | | |  |
| Crèche 2 days a week # Grandparents support - when necessary | |  |  | | | 1.49^*^ |
|  | |  |  | | | [1.02,2.16] |
| Crèche 5 days a week # Grandparents support - sometimes | |  |  | | | 0.55^*^ |
|  | |  |  | | | [0.32,0.95] |
| Crèche 5 days a week # Grandparents support – working days | |  |  | | | 0.54^*^ |
|  | |  |  | | | [0.33,0.90] |
| Crèche 5 days a week # Grandparents support – Everyday | |  |  | | | 0.53^*^ |
|  | |  |  | | | [0.33,0.87] |
| ***Interaction effects with feasible number of children*** | |  |  | | |  |
| Paternity leave of 98 days # 2 children realistically | |  |  | | | 1.43^*^ |
|  | |  |  | | | [1.08,1.88] |
| ***Interaction effects with ideal number of children*** | |  |  | | |  |
| Paternity leave of 20 days # 2 children ideally | |  |  | | | 1.57^**^ |
|  | |  |  | | | [1.20,2.06] |
| ***N*** | 12060 | | | 12060 | 12060 | |
| Reported estimates are odds ratio, with 95% confidence interval in the squared bracket below.  ^*^ *p* < 0.05, ^**^ *p* < 0.01, ^***^ *p* < 0.001 | | | | | | |

**Appendix F: Heterogeneous Preferences for Family Policies in Terms of the Decision to Have the First, Second and Third Child**

To explore whether women’s preferences for family formation support policies differ by whether they were considering having their first, second, or third child, equation (1) was re-estimated on two additional subsamples from the original data: women who had no children and women who already had two children. Results are reported in Table A4. As can be seen, preferences were remarkably similar across the groups. Only women considering their first child demonstrated a stronger preference for career progression protection and extended paternity leave, while showing a lower preference for extended maternity leave than women considering whether to have a second child. This shows that our conclusions about the impact of policies on fertility intention is robust to whether women are considering having their first, second, or third child.

| **Table A4: Estimated Heterogeneous Preferences When Considering to Have The First, Second and Third Child** | | | |
| --- | --- | --- | --- |
|  | Column A | Column B | Column C |
|  | Base Model -  Equation (1) | Intention of having the first child | Intention of having the third child |
| ***Baby stipend (effect in 100s of CNY)*** | 1.23^***^ | 1.20^***^ | 1.27^***^ |
|  | [1.22,1.25] | [1.17,1.24] | [1.23,1.30] |
| ***Paid maternity leave*** | *Reference Level* | | |
| 98 days |  |  |  |
| 158 days | 1.92^***^ | 1.70^***^ | 1.91^***^ |
|  | [1.74,2.12] | [1.38,2.10] | [1.64,2.23] |
| 270 days | 3.96^***^ | 3.61^***^ | 3.34^***^ |
|  | [3.44,4.56] | [2.69,4.85] | [2.67,4.19] |
| ***Paid paternity leave*** | *Reference Level* | | |
| None |  |  |  |
| 20 days | 2.60^***^ | 2.83^***^ | 2.42^***^ |
|  | [2.35,2.87] | [2.25,3.56] | [2.06,2.85] |
| 98 days | 3.49^***^ | 4.48^***^ | 3.14^***^ |
|  | [3.11,3.91] | [3.51,5.72] | [2.65,3.73] |
| ***Childcare support*** | *Reference Level* | | |
| None |  |  |  |
| Free access to crèche 2 days a week for 3 years | 1.62^***^ | 2.00^***^ | 1.70^***^ |
|  | [1.48,1.78] | [1.66,2.40] | [1.48,1.96] |
| Free access to crèche 5 days a week for 3 years | 2.55^***^ | 2.45^***^ | 2.44^***^ |
|  | [2.24,2.90] | [1.94,3.09] | [1.97,3.01] |
| ***Career progression*** | *Reference Level* | | |
| Promotion delayed by 2 years |  |  |  |
| Promotion delayed by 1 year | 1.55^***^ | 1.65^***^ | 1.19^*^ |
|  | [1.40,1.70] | [1.33,2.05] | [1.03,1.38] |
| Promotion not affected | 4.76^***^ | 5.34^***^ | 3.08^***^ |
|  | [4.16,5.45] | [3.98,7.18] | [2.55,3.73] |
| ***N*** | 1005 | 230 | 432 |
| Reported estimates are odds ratio, with 95% confidence interval in the squared bracket below.  ^*^ *p* < 0.05, ^**^ *p* < 0.01, ^***^ *p* < 0.001 | | | |

**References**

Binaku-Hajrullahu, L. (2019). Maternity Leaves and Their Effects on Career and Career Development.

Bridges, J. F., Hauber, A. B., Marshall, D., Lloyd, A., Prosser, L. A., Regier, D. A., Johnson, F. R., & Mauskopf, J. (2011). Conjoint analysis applications in health—a checklist: a report of the ISPOR Good Research Practices for Conjoint Analysis Task Force. *Value in Health*, *14*(4), 403-413.

Choi, S.-w., Yellow Horse, A. J., & Yang, T.-C. (2018). Family policies and working women’s fertility intentions in South Korea. *Asian Population Studies*, *14*(3), 251-270.

Jing, W., Liu, J., Ma, Q., Zhang, S., Li, Y., & Liu, M. (2022). Fertility intentions to have a second or third child under China’s three-child policy: a national cross-sectional study. *Human Reproduction*, *37*(8), 1907-1918.

Jones, G. W. (2019). Ultra-low fertility in East Asia: policy responses and challenges. *Asian Population Studies*, *15*(2), 131-149. <https://doi.org/10.1080/17441730.2019.1594656>

Li, M., Niu, D., Lin, Y., Li, S., & Wang, X. (2024). Research on the effectiveness of second-child subsidy policy in high-cold areas based on multi-level model. *Journal of Education, Humanities and Social Sciences*, *35*. <https://doi.org/10.54097/4v7bj093>

Staehelin, K., Bertea, P. C., & Stutz, E. Z. (2007). Length of maternity leave and health of mother and child – a review. *International Journal of Public Health*, *52*(4), 202-209. <https://doi.org/10.1007/s00038-007-5122-1>

World Population Review. (2024). *Maternity leave by country*. Retrieved 23 November from <https://worldpopulationreview.com/country-rankings/maternity-leave-by-country>
